# Deriving Greater Value from Malaria Bed Nets through Extending Net Retention: A Modelling Study

**DOI:** 10.1101/2025.08.02.25332507

**Authors:** Jessica Floyd, Anna Trett, Monica Golumbeanu, Clara Champagne, Abby Ward, Emilie Pothin, Justin Cohen, Tara Seethaler

## Abstract

Insecticide-treated nets (ITNs) are central to malaria prevention in sub-Saharan Africa, but their effectiveness depends on how long households retain and use them. Although ITNs are designed to last three years, recent evidence suggests median retention times are significantly shorter in most countries, creating gaps in protection between mass distribution campaigns. In this study, we model the potential impact of improving ITN retention by enhancing physical durability on malaria burden across 43 countries in sub-Saharan Africa using an individual-based malaria transmission model. We simulate scenarios in which ITN retention times are extended by 1 to 8 months beyond current country-specific baselines and compare averted cases relative to a business-as-usual scenario. Results show that a 4-month increase in retention could avert 6% of malaria cases over three years; and that to get the same amount of impact from increasing ITN access would require an additional 171 million nets. Gains from improved retention are greatest in the first few months of extension and accumulate over time, making durability enhancements a cost-effective strategy for maintaining protection as funding constraints grow. These findings support the integration of predictive durability metrics, such as the Resistance to Damage (RD) score, into ITN procurement decisions and highlights the importance of selecting well performing, high-value ITNs to strengthen malaria control without deploying more ITNs.

## Introduction

Insecticide-treated nets (ITNs) are a vital tool in the fight against malaria in sub-Saharan Africa, where the disease remains a leading cause of illness and death^1^. Distribution campaigns and public health initiatives have seen the distribution of over 3.4 billion ITNs since 2004, contributing significantly to the reduction of global malaria over the past two decades^2^. ITNs are impregnated with insecticides like pyrethroids, creating a physical and chemical barrier that protects individuals from mosquito bites, particularly at night when *Anopheles* mosquitoes, the primary vector of malaria, are most active. ITNs not only reduce direct human-mosquito contact, but also lower the local mosquito population and transmission rates through community-wide usage^3,4^.

Mass distribution campaigns have primarily and historically been conducted every three years, in line with the expected lifespan of ITNs^5,6^. The duration for which households continue to use and maintain ITNs - known as the retention time - is critical to the cost-effectiveness of ITNs as a malaria intervention. A 2021 study estimated that the median retention time for ITNs in Africa was approximately 1.64 years, with an interquartile range of 1.33 to 2.37 years^7^. Notably, 35 out of the 40 countries studied exhibited median retention times of less than 3 years, suggesting that ITNs are often discarded more quickly than the commonly assumed lifespan of 3 years^8,9^. As such, many areas face severe coverage gaps in protection between campaign cycles^10^.

Recent research has demonstrated that the physical durability of ITNs strongly influences their retention and functional lifespan in the field, independent of mosquito susceptibility to insecticide treatment^11–13^. This led to the development of the Resistance to Damage (RD) score developed by the Nonwovens Institute Research Initiative (NIRI) – a quantitative, standardized, lab-collected metric that predicts a net’s ability to withstand wear and tear^14,15^. Further refinement of the RD score has increased its predictiveness for ITN survival, with a recent study noting that an increase in the RD score from the lowest score of the current range of products to the highest was associated with 13 extra months of ITN lifespan in the field^16^.

With recent reductions to global malaria donor funding, maximizing the impact of ITN distribution has become more critical than ever. Extending net lifespan could reduce the coverage gaps between mass distribution campaigns, increase the protection between three year campaign cycles, and encourage higher usage. In this study, we use an agent-based model of *P. falciparum* malaria transmission to estimate the potential impact of improving ITN retention on malaria cases across sub-Saharan Africa and compared the outputs to increasing ITN usage under different scenarios. Our findings quantify the relative benefits of longer ITN retention compared to increasing ITN procurement volumes and highlight the potential value of incorporating predictive durability metrics, such as the RD score, into procurement decisions.

## Methodology

To translate increases in average ITN RD scores and resulting additional months of ITN lifespan into prolonged malaria protection, we modelled malaria transmission based on increased ITN retention times. To estimate the impact on malaria transmission and burden, we used OpenMalaria, an open-source, individual-based model of *P. falciparum* malaria transmission^17^. OpenMalaria provides a robust framework for simulating malaria epidemiology and control interventions under diverse settings. Our modelling approach focused on varying ITN retention times and, separately, ITN ‘initial usage’ rates based on country-specific estimates. We define ITN retention as the duration that households retain ITNs after initial distribution and ITN initial usage as the proportion of the population sleeping under an ITN at the beginning of the campaign. We modelled the impact of a single ITN distribution conducted at the beginning of 2023 on malaria cases in sub-Saharan Africa over the subsequent three years. We used 2023 as a starting point as the model was calibrated to historical data available through the end of 2022. To systematically evaluate the impact of increasing ITN retention times and initial usage rates, we compared malaria cases averted under scenarios with incrementally increased retention (1–8 months) or initial usage (1%–8%) to a baseline where retention and initial usage remained constant. Insecticide efficacy decayed exponentially, which was parameterised using data from experimental hut trials^18,19^ and this rate of decay was held constant across simulations.

### Model calibration

We first calibrated the model with the aim of reproducing historical *P. falciparum* prevalence estimates provided by the Malaria Atlas Project (MAP) in each setting (defined as the first administrative level) for all 43 malaria-endemic countries in sub-Saharan Africa. The model was calibrated using historical national and sub-national estimates of malaria, prevalence, vector composition and behaviour, seasonality, population and historical intervention coverages, which enabled us to create simulations tailored to the epidemiologic profile of each country and the specific data used and their sources are given in Table 1. We used these country profiles to run simulations and calibrate with a profile likelihood-based approach which retrieves the best-fitting entomological inoculation rate (EIR) and associated uncertainty bounds^20^. The simulated transmission intensity which best matched the observed malaria prevalence according to MAP estimates was identified and used for future simulations.

**Table 1:** Data and sources used to parameterise OpenMalaria during calibration and future simulations.

| Parameter | Details | Reference |
| --- | --- | --- |
| *P. falciparum* prevalence | Admin-1 level prevalence estimates provided by MAP | 21,22 |
| Vector composition (for *Anopheles gambiae*, *funestus* and *arabiensis*) | Relative abundances of vector species at country level | 23 |
| Vector behaviour (for *Anopheles gambiae*, *funestus* and *arabiensis*) | Proportions of bites occurring outdoors/indoors: calculated at country level using the AnophelesModel R package | 18,24 |
| Seasonality | Monthly precipitation aggregated to Admin-1 from WorldClim | 24,25 |
| Population | Admin-1 level population estimates provided by MAP | 24 |
| Age structure | Proportions of population in 5-year age brackets by admin-1 | 24,26 |
| ITN usage | Admin-1 estimates of ITN usage provided by MAP | 24 |
| ITN efficacy | Parameterised from experimental hut trials and uses a standard pyrethroid ITN in the presence of moderate pyrethroid resistance | 27 |
| IRS coverage | Admin-1 estimates of IRS coverage provided by MAP | 24 |
| IRS efficacy | Parameterised from experimental hut trials | 19 |
| Effective treatment coverage | Estimates of the proportion of the population effectively treated for malaria, provided by MAP | 24 |
| SMC coverage | Estimates of the proportion of the under-5 population receiving SMC, provided by MAP | 24 |
| SMC efficacy | 25 days blood clearance | 28 |
| Importation rate | 10 infections per 1000 people per year in each commune | 29 |

### Simulations

In the future simulations, for 582 settings in the 43 countries we ran 5 stochastic replicates for each setting and three values of EIR (the central estimate and the upper and lower uncertainty bounds) for each of the 9 scenarios, giving a total of 78,570 simulations. To reduce the computational resources required to run these simulations, we gave setting a population size of 9,000, then rescaled the results of the simulations up to the true population size of the setting using estimates given in Table 1 during post-processing.

### Scenarios

We modelled changes in ITN retention time by modifying the attrition of ITNs in the ITN parameterisation for OpenMalaria^30,31^. ITN attrition is modelled using a *smooth-compact* decay curve, a mathematical function that describes how ITNs are discarded by users over time, with a half-life representing the time at which 50% of ITNs have been discarded by users, or the median retention time. This approach allowed us to simulate how long households retain ITNs before they are discarded due to factors like wear and tear, loss, or reduced perceived utility. The decay curve provides a continuous representation of retention dynamics, transitioning smoothly from high retention levels immediately after distribution to progressively lower levels over the modelled time horizon, and is represented by the following equation:

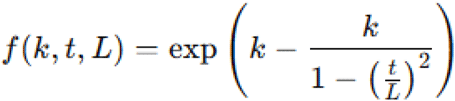

Where:

*k* is a shape parameter with no dimension,

*t* is the time elapsed since the ITN distribution, and

*L* is the maximum ITN retention time.

An example of the ITN retention time decay curves used in the modelling for a single country is given in Figure 1:

**Figure 1.**
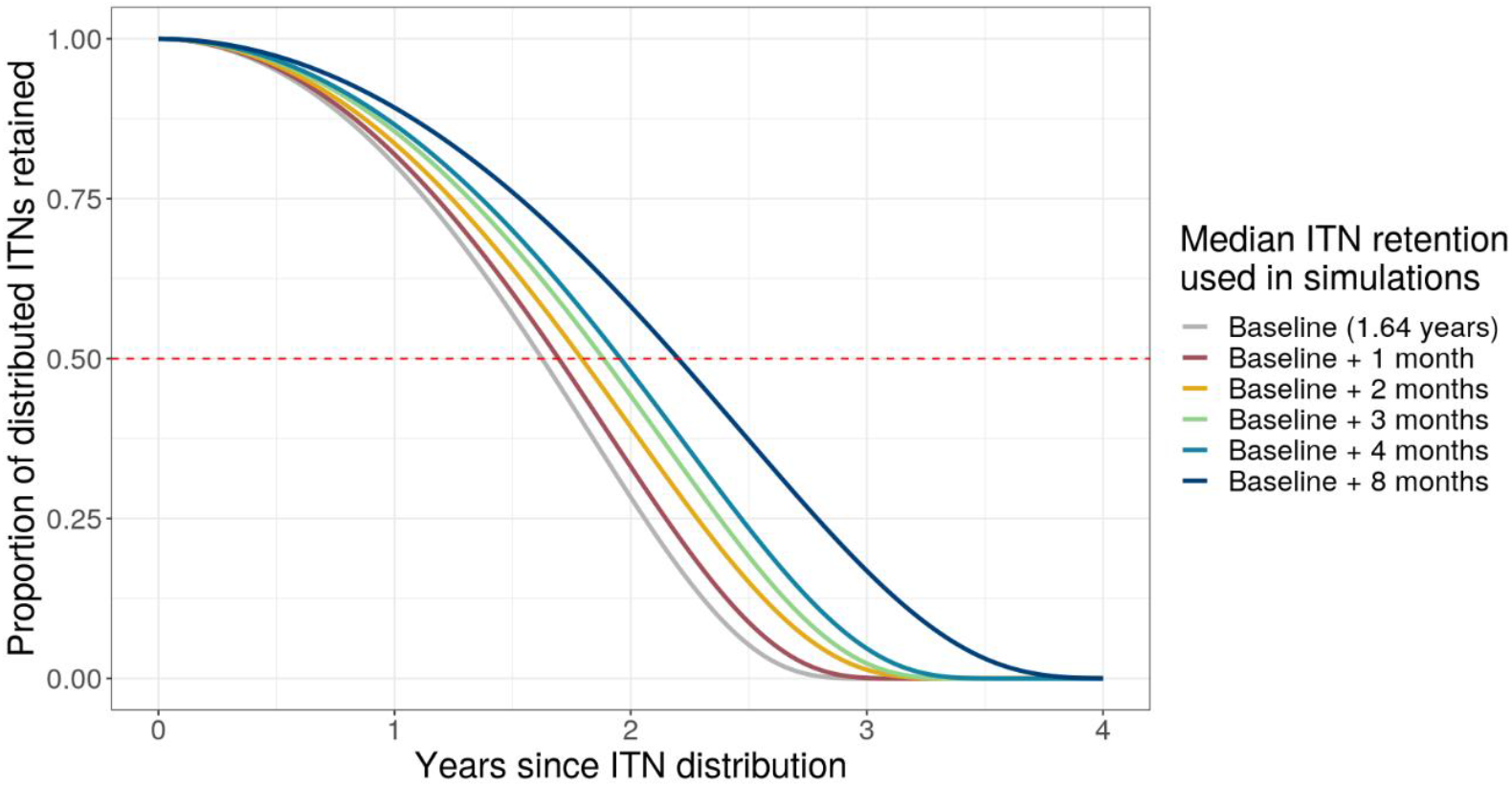
Example of modelled ITN retention time decay curves for a single given median retention time of 1.64 years (baseline for this example), plus an additional 1, 2, 3, 4 and 8 months. The median retention time (at which 50% of the population have discarded their ITN) is shown by the red dashed line.

In the baseline scenario, we used country-specific median ITN retention times as reported in Bertozzi-Villa et al.^7^, which provides empirical estimates of ITN ownership, retention times and usage decay over time. Supplementary Table 1 shows the baseline country-specific median ITN retention times used in the simulations. ITN initial usage rates in the baseline scenario were set at the highest ITN usage level observed at the first administrative level in the past three years according to estimates provided by MAP^24^. In the alternative scenarios, we either extended median retention times incrementally by 1, 2, 3, 4 and 8 months, or increased initial usage rates of 1%, 2%, 3%, 4%, 5%, 6%, 7% and 8% to observe the impact on malaria cases averted compared to baseline. In the future simulations of extended retention times, we ensured that all other parameters, such as ITN usage at the beginning of the campaign (‘initial usage’) and malaria case management interventions, were held constant to isolate the effect of changes in retention time. Retention time increases in all scenarios were constrained to ensure that median retention time did not exceed three years in countries that had median retention times of under three years at baseline. Similarly, in the future simulations of increased ITN initial usage rates, we ensured that all other parameters such as ITN retention rates and case management interventions were held constant and ITN initial usage was constrained to 100% in all scenarios.

### Modelling outputs

For all scenarios modelled, we calculated the cumulative percentage of malaria cases averted by comparing mean malaria cases in the baseline and extended ITN retention and ITN initial usage scenarios, expressed as a percentage reduction summed over three years post-distribution:

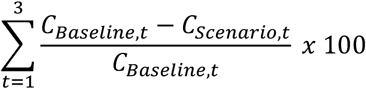

Where:

*C*_*Baseline,t*_ = mean malaria cases in year *t* under baseline retention and initial usage rates and

*C*_*Scenario,t*_ = mean malaria cases in year *t* under extended retention or increased initial usage rates

This approach allowed us to quantify the potential malaria cases averted in sub-Saharan Africa by interventions aimed at improving ITN longevity, such as enhancing the physical durability of ITNs, and by interventions aimed at increasing the proportion of the population using an ITN.

### Estimating the ITN supply required to increase usage rates

Lastly, we used estimates from the Malaria Atlas Project’s ITN stock-and-flow model^7^ to predict the net supply required to achieve the target ITN initial usage levels in each scenario. This model accounts for ITN distribution, access, usage and decay in retention over time, allowing us to translate desired changes in initial usage into corresponding ITN procurement volumes. We downloaded estimates from the ITN dashboard^24^ produced by MAP and for each first-level admin-1 across all countries included in the analysis, we fitted Generalised Additive Models (GAMs) to estimate the relationship between the number of ITNs per capita and observed initial usage rates. These flexible models allowed us to interpolate and predict the per capita ITN supply needed to achieve the initial usage levels implemented in each modelling scenario. We then scaled up the ITN-per-capita rates to the size of the population in each admin-1 unit and summed over the 43 countries to get total numbers of ITNs needed in each scenario.

## Results

### Extending ITN retention times by 4 months could avert 6% of malaria cases in sub-Saharan Africa

Across all modelled scenarios and country settings, increasing ITN retention times resulted in reductions in malaria cases. Here we present the cumulative percentage of cases averted by increasing retention times.

As shown in Figure 2, incremental increases in ITN retention time - from one to eight months beyond baseline country-specific estimates of retention - led to progressively more cases averted over the three-year post-ITN distribution period. Extending ITN retention times by two, four and eight months resulted in cumulative percentages of malaria cases averted of 3.43% (CIs:2.68% - 4.23%), 6.34% (CIs:5.14% - 7.59%) and 10.9% (CIs:9.0% - 12.9%), respectively over the three years and shown in the third set of bars in Figure 2. The relationship between retention and cases averted was non-linear, with the largest marginal gains observed in the first few months of extension and this relationship is more pronounced within the third year following ITN distribution.

**Figure 2.**
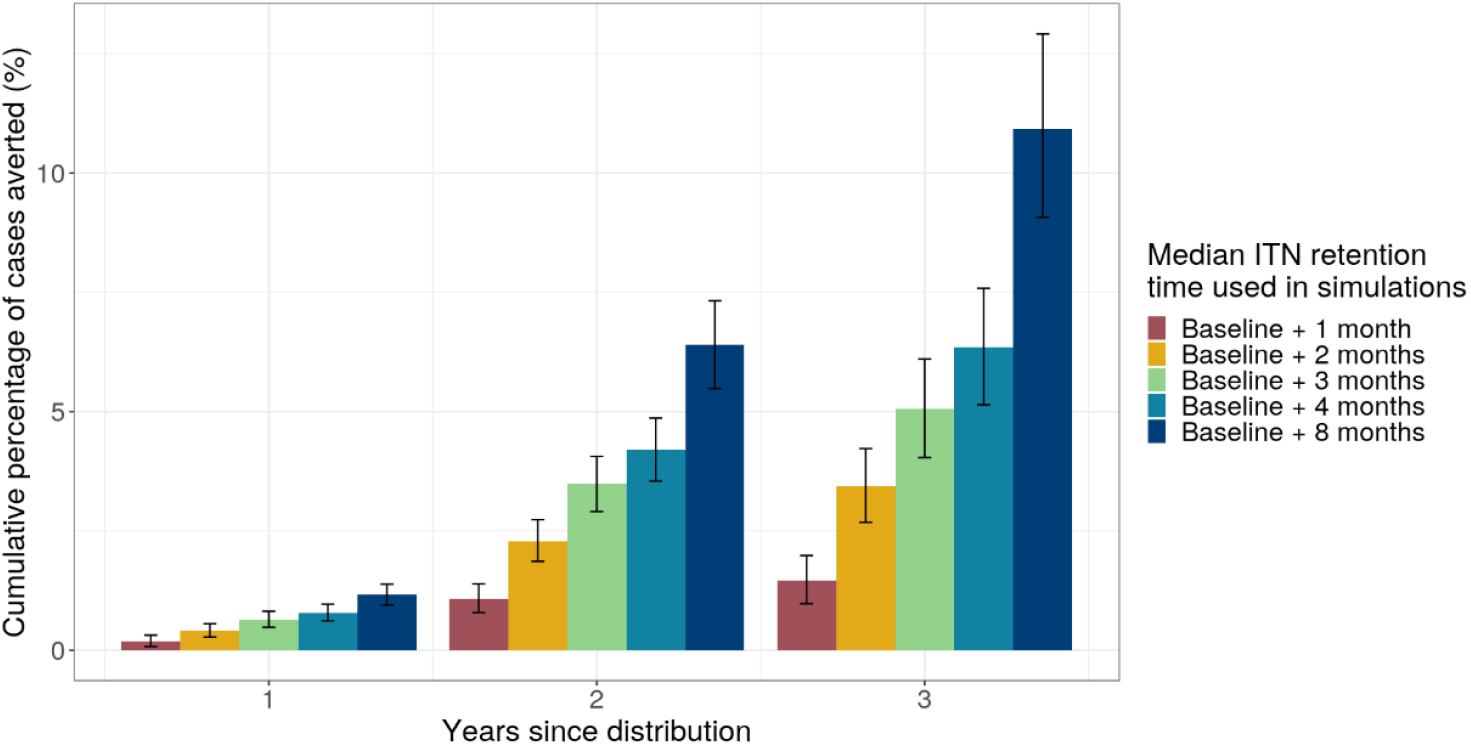
The cumulative percentage of malaria cases averted by extending ITN retention times by 1, 2, 3, 4 and 8 months compared to baseline ITN retention times. These are the total cumulative percentages of cases averted across settings in sub-Saharan Africa at the end of each year after the initial deployment.

### Increases in ITN retention times by 4 months through improved ITN lifespan could provide similar impact to increasing ITN initial usage by 7%

To contextualise the epidemiological impact of increased ITN retention, we compared it to the impact of increased ITN initial usage (defined as the proportion of the population that slept under an ITN the previous night at the beginning of the campaign) (Figure 3). Our findings show that even modest increases in ITN retention - ranging from 2 to 4 months - could avert as many malaria cases as a 4-7% increase in ITN initial usage. For instance, in simulations where ITN initial usage was increased by 7%, extending the retention period by just 4 months resulted in similar levels of malaria case reduction over a three-year distribution cycle. Notably, increased initial usage has an immediate impact within the first year after distribution, whereas extending net lifespan does not significantly affect malaria cases in the first year, since most nets already remain effective during that period and the added benefit only begins after the first year (Supplementary Figure 1).

**Figure 3.**
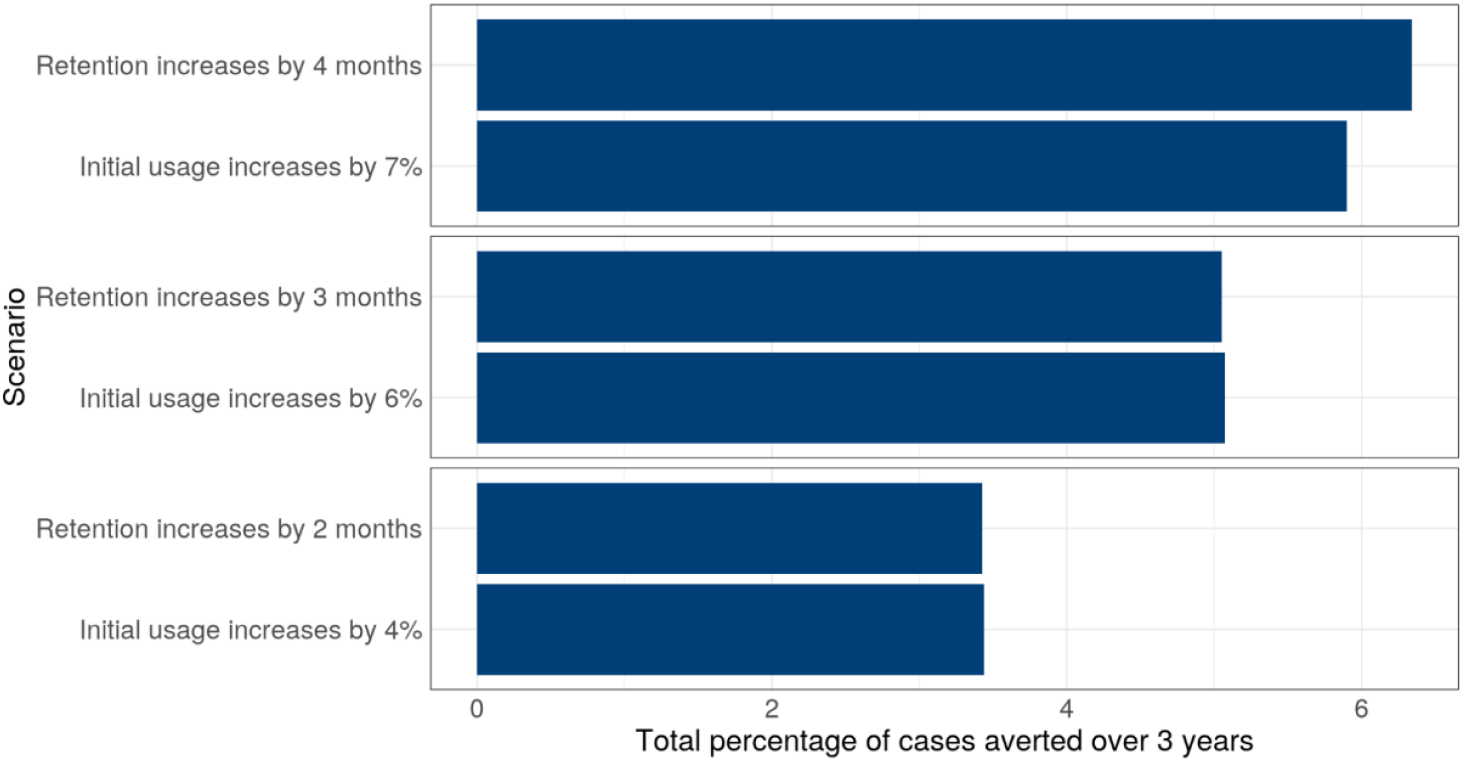
Selected ITN retention and initial usage scenarios with the nearest equivalent overall impact. In each scenario, mean percentages of malaria episodes averted by increased retention or initial usage compared to baseline ITN retention and initial usage are given. In the retention scenarios and initial usage scenarios, the baselines are the country-specific median retention time and the maximum ITN usage observed in last 3 years, respectively.

### A 7% increase in ITN initial usage would require distributing an additional 171 million ITNs, meaning that achieving the equivalent impact of a 4-month increase in retention could increase the unit value of ITNs by approximately USD$1.35

Given current global funding constraints, we estimated the number of additional nets that would need to be distributed to achieve the same epidemiological outcome as extending retention time. Applying the above finding that 4 months retention equates to 7% increase in ITN initial usage in terms of epidemiological impact, we then applied the MAP stock-and-flow model to show that a 7% initial usage increase would require 171 million additional ITNs to be distributed across the 582 settings in 43 countries (Table 2).

**Table 2:**
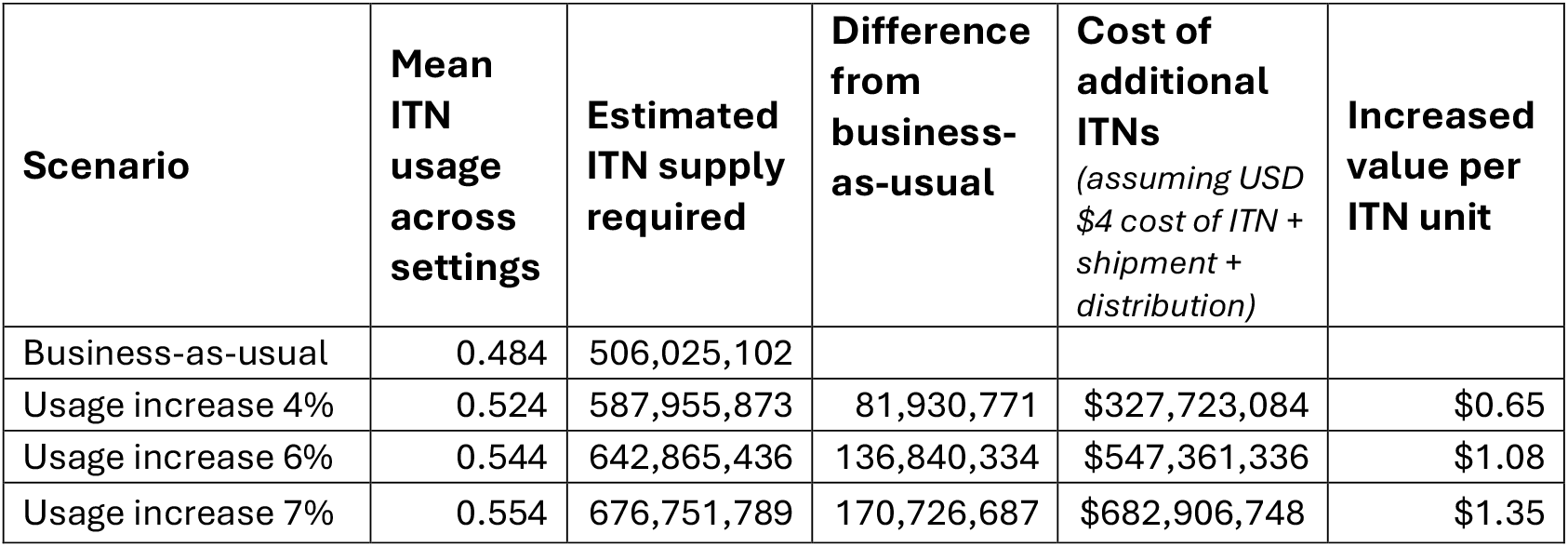
Mean ITN initial usage, ITN supply required to achieve that usage, and a costed example of the increased value of longer-lasting ITNs compared to business-as-usual in each scenario. The increased value is calculated as the cost of procuring and distributing the additional ITNs (using an example total cost of USD$4 per ITN distributed) divided by the number of ITNs required in the business-as-usual scenario.

| **Scenario** | **Mean ITN usage across settings** | **Estimated ITN supply required** | **Difference from business-as-usual** | **Cost of additional ITNs**<br>*(assuming USD \$4 cost of ITN + shipment + distribution)* | **Increased value per ITN unit** |
| --- | --- | --- | --- | --- | --- |
| Business-as-usual | 0.484 | 506,025,102 |  |  |  |
| Usage increase 4% | 0.524 | 587,955,873 | 81,930,771 | \$327,723,084 | \$0.65 |
| Usage increase 6% | 0.544 | 642,865,436 | 136,840,334 | \$547,361,336 | \$1.08 |
| Usage increase 7% | 0.554 | 676,751,789 | 170,726,687 | \$682,906,748 | \$1.35 |

To explore the economic implications, we conducted an illustrative costing analysis to address the question: “If procurers continue to purchase the same number of ITNs, how much more should they be willing to pay per unit for a longer-lasting ITN?”. We assumed a conservative baseline cost of USD$4 per ITN, including commodity procurement, shipment, and distribution. Using this assumption, we calculated the total cost of additional ITNs required under each increased initial usage scenario. We then divided these costs by the number of ITNs in the business-as-usual scenario, yielding an indicative estimate of the added value per ITN if those ITNs could deliver the same impact on malaria cases as the higher initial usage scenarios.

## Discussion

Our findings demonstrate that increasing ITN retention times, even by a few months, could substantially reduce malaria burden across diverse transmission settings in sub-Saharan Africa. Incremental improvements in retention-of just a few months beyond current baseline estimates - produced non-linear health gains, with up to 6% of malaria cases averted with 4 months extra retention. This highlights the significant potential of modest enhancements in ITN lifespan especially in contexts where programmatic or financial constraints limit the feasibility of widespread ITN campaigns.

Although it is well known that increased ITN durability has been associated with reduced malaria outcomes, these findings underscore the potential of improving ITN retention as a viable and efficient complement to coverage-focused strategies. A wealth of previous evidence has shown that increasing coverage and consistent use of ITNs substantially reduces malaria infection rates and child mortality, both at the individual and community level^32^. Additionally, previous modelling looking at coverage levels required to control malaria for varying ITN efficacies concluded that ITNs with longer lifespans performed better than those with shorter lifespans^33^.

While increasing ITN initial usage remains a powerful lever for malaria control, the financial and operational demands of distributing over 170 million additional nets to achieve a 7% usage increase are substantial. In contrast, extending net retention by a few months through procurement of more durable products that are currently on the market may offer a similarly impactful alternative with fewer logistical burdens and less impact on the environment through reducing the need for plastic production. This is particularly relevant in settings where frequent net replacement is difficult to sustain.

Moreover, the temporal dynamics of these interventions are crucial for programmatic planning. While increased initial usage leads to immediate reductions in malaria burden, gains from improved retention accrue more gradually as they delay the decay in protection between campaigns. This delayed but sustained benefit makes retention improvements especially valuable for preserving coverage in the later years of a distribution cycle, when many areas face sharp declines in effective ITN access. In settings where funding cuts or supply chain constraints limit the feasibility of frequent mass campaigns, extending net longevity can act as a strategic buffer.

Together, these insights highlight the importance of optimizing the value of ITNs distributed, with a greater emphasis on durability as a cost-effective pathway to maintaining protection in vulnerable populations.

Several limitations must be considered when interpreting these results. First, we applied retention time improvements and initial usage reductions uniformly across all settings, despite likely variation in local behaviours, ITN product selection, and operational conditions. Second, we modelled a single type of ITN, thus assuming that ITNs deployed in the future would retain similar efficacy to those currently in use, and not accounting for the potential impact of the growing shift to dual AI ITNs in response to resistance. This also omits consideration of insecticide resistance, which could reduce the effective lifespan of nets in ways not captured by our retention metric.

Third, we modelled only a single ITN distribution occurring at the beginning of 2023 for all countries, without accounting for the seasonal timing of actual campaign schedules or the effects of continuous distribution strategies (e.g., through schools or health facilities). These additional layers of distribution could interact with net durability in complex ways - either reinforcing or dampening the gains seen in our simulations. Moreover, we assumed a maximum operational lifespan of three years for ITNs; thus, settings where retention times already exceed this threshold were not expected to benefit further from additional improvements.

Lastly, our analysis was conducted at the first administrative level due to data availability constraints and the significant computational demand of higher-resolution modelling. While this enabled a broad comparative analysis across settings, it obscures finer-scale heterogeneity that could influence both ITN effectiveness and the practical value of durability improvements.

Despite these limitations, our results support the strategic value of enhancing ITN physical durability, particularly in environments where sustaining high coverage is logistically or financially constrained. Policymakers and funding agencies may consider durability as a key consideration in ITN procurement and innovation, recognising that links closely to retention and even small gains in retention can help sustain protection and mitigate emerging threats to malaria control.

### Conclusion and Policy Implications

As global malaria funding faces increasing pressure, maximizing the impact from existing investments becomes more critical than ever. This study demonstrates that modest extensions in ITN retention time can yield substantial reductions in malaria cases—sometimes rivalling the benefits of considerably increasing net usage levels. Procurers may enhance the long-term value of distributed ITNs by incorporating predictive durability metrics, such as the RD score, into procurement decisions. Procurement decisionmakers can also use these estimates when ascribing value to nets with expected longer lifespans. In doing so, they may help create market incentives for the design and production of more durable ITNs. National malaria programs and global funders should consider retention-focused strategies as a cost-effective complement to distribution campaigns, particularly in countries where ITNs are currently discarded well before their intended three-year lifespan. Investing in net durability may not only improve health outcomes but also enhance the efficiency and sustainability of malaria control programs.

## Data Availability

All data produced in the present study are available upon reasonable request to the authors.

## Acknowledgements

The authors would like to thank Steve Isaacs, Jeanne Lemant, Christian Selinger, Amelia Bertozzi-Villa and Kate Kolazinski for valuable discussions. Calculations were performed at sciCORE (http://scicore.unibas.ch/) scientific computing centre at the University of Basel (Basel, Switzerland).

## References

1. World Malaria Report 2023. (2023).

2. J Weiss, A. D. et al. Mapping the global prevalence, incidence, and mortality of Plasmodium falciparum and Plasmodium vivax malaria, 2000–22: a spatial and temporal modelling study. The Lancet 405, 979–990 (2025).

3. Binka, F. N. et al. Impact of permethrin impregnated bednets on child mortality in Kassena-Nankana district, Ghana: A randomized controlled trial. Tropical Medicine and International Health 1, 147–154 (1996).

4. Hawley, W. A. et al. Community-wide effects of permethrin-treated bed nets on child mortality and malaria morbidity in western Kenya. American Journal of Tropical Medicine and Hygiene 68, 121–127 (2003).

5. WHO guidance note for estimating the longevity of long-lasting insecticidal nets in malaria control September 2013 | WHO | Regional Office for Africa. https://www.afro.who.int/publications/who-guidance-note-estimating-longevity-long-lasting-insecticidal-nets-malaria-control.

6. World Health Organization. Guidelines for laboratory and field-testing of long-lasting insecticidal nets. Who/Htm/Ntd/Whopes/2013.1 93 (2013).

7. Bertozzi-Villa, A. et al. Maps and metrics of insecticide-treated net access, use, and nets-per-capita in Africa from 2000-2020. Nature Communications 2021 12:1 12, 1–12 (2021).

8. Raharinjatovo, J. et al. Physical and insecticidal durability of Interceptor^®^, Interceptor^®^ G2, and PermaNet^®^ 3.0 insecticide-treated nets in Burkina Faso: results of durability monitoring in three sites from 2019 to 2022. Malar J 23, 1–19 (2024).

9. Ngufor, C. et al. The attrition, physical and insecticidal durability of two dual active ingredient nets (Interceptor^®^ G2 and Royal Guard^®^) in Benin, West Africa: results from a durability study embedded in a cluster randomised controlled trial. Parasit Vectors 17, 420 (2024).

10. Paaijmans, K. P. & Lobo, N. F. Gaps in protection: the actual challenge in malaria elimination. Malar J 22, 1–4 (2023).

11. Massue, D. J. et al. Durability of Olyset campaign nets distributed between 2009 and 2011 in eight districts of Tanzania. Malar J 15, 1–11 (2016).

12. Koenker, H. et al. What happens to lost nets: A multi-country analysis of reasons for LLIN attrition using 14 household surveys in four countries. Malar J 13, 1–10 (2014).

13. Hiruy, H. N. et al. The effect of long-lasting insecticidal nets (LLINs) physical integrity on utilization. Malar J 20, 1–10 (2021).

14. Kilian, A. et al. Correlation of textile ‘resistance to damage’ scores with actual physical survival of long-lasting insecticidal nets in the field. Malar J 20, 1–10 (2021).

15. Wheldrake, A., Guillemois, E., Chetty, V., Kilian, A. & Russell, S. J. Development of a single resistance to damage metric for mosquito nets related to physical integrity in the field. Malar J 20, 1–13 (2021).

16. Mechan, F. et al. Refining the resistance-to-damage (RD) score to predict operational Insecticide-Treated Net lifespan and identify paths to innovation. bioRxiv 2025.03.02.641027 (2025) doi:10.1101/2025.03.02.641027.

17. Smith, T. et al. Ensemble Modeling of the Likely Public Health Impact of a Pre-Erythrocytic Malaria Vaccine. PLoS Med 9, e1001157 (2012).

18. Golumbeanu, M. et al. AnophelesModel: An R package to interface mosquito bionomics, human exposure and intervention effects with models of malaria intervention impact. PLoS Comput Biol 20, e1011609 (2024).

19. Tchicaya, E. S. et al. Micro-encapsulated pirimiphos-methyl shows high insecticidal efficacy and long residual activity against pyrethroid-resistant malaria vectors in central Côte d’Ivoire. Malar J 13, 332 (2014).

20. Ionides, E. L., Breto, C., Park, J., Smith, R. A. & King, A. A. Monte Carlo profile confidence intervals for dynamic systems. J R Soc Interface 14, (2017).

21. Weiss, D. J. et al. Mapping the global prevalence, incidence, and mortality of Plasmodium falciparum and Plasmodium vivax malaria, 2000–22: a spatial and temporal modelling study. The Lancet 405, 979–990 (2025).

22. Weiss, D. J. et al. Mapping the global prevalence, incidence, and mortality of Plasmodium falciparum, 2000–17: a spatial and temporal modelling study. Lancet 394, 322 (2019).

23. Sinka, M. E. et al. Modelling the relative abundance of the primary African vectors of malaria before and after the implementation of indoor, insecticide-based vector control. Malar J 15, 1–10 (2016).

24. Malaria Atlas Project | Data. https://data.malariaatlas.org/trends?year=2022&metricGroup=Malaria&geographicLevel=admin0&metricSubcategory=Pf&metricType=rate&metricName=incidence.

25. Fick, S. E. & Hijmans, R. J. WorldClim 2: new 1-km spatial resolution climate surfaces for global land areas. International Journal of Climatology 37, 4302–4315 (2017).

26. WorldPop :: Age and sex structures. https://hub.worldpop.org/geodata/listing?id=65.

27. Briët, O. J. T. et al. Effects of pyrethroid resistance on the cost effectiveness of a mass distribution of long-lasting insecticidal nets: a modelling study. Malar J 12, 77 (2013).

28. Chandramohan, D. et al. Effect of Adding Azithromycin to Seasonal Malaria Chemoprevention. New England Journal of Medicine 380, 2197–2206 (2019).

29. Tatem, A. J. et al. The use of mobile phone data for the estimation of the travel patterns and imported Plasmodium falciparum rates among Zanzibar residents. Malar J 8, 287 (2009).

30. OpenMalaria Schema (latest version). https://swisstph.github.io/openmalaria/schema-latest.html#elt-attritionOfNets (2025).

31. Home · SwissTPH/openmalaria Wiki · GitHub. https://github.com/SwissTPH/openmalaria/wiki (2025).

32. Pryce, J., Richardson, M. & Lengeler, C. Insecticide-treated nets for preventing malaria. Cochrane Database of Systematic Reviews 2018, (2018).

33. Ngonghala, C. N., Del Valle, S. Y., Zhao, R. & Mohammed-Awel, J. Quantifying the impact of decay in bed-net efficacy on malaria transmission. J Theor Biol 363, 247–261 (2014).

