## Supplementary Information for "Deriving Greater Value from Malaria Bed Nets through Extending Net Retention: A Modelling Study"

### Supplementary Information for the Manuscript "Deriving Greater Value from Malaria Bed Nets through Extending Net Retention: A Modelling Study"

*Supplementary table 1: Median ITN retention rates from Bertozzi-Villa et. al.[15] and used as baseline country-specific median ITN retention times in simulations. The confidence intervals around the median retention times are given in brackets and the number of Demographic Health Surveys/Malaria Indicator Surveys (DHS/MIS) that were used to fit each country in the Bertozzi-Villa model is also listed.*

| Country | Median Retention Time (Years) | Survey Count |
|---|---|---|
| Angola | 1.10 (1.01, 1.26) | 3 |
| Benin | 1.07 (1.01, 1.17) | 4 |
| Burkina Faso | 1.58 (1.41, 1.76) | 5 |
| Burundi | 1.31 (1.14, 1.47) | 4 |
| Cameroon | 3.49 (3.24, 3.78) | 4 |
| Central African Republic | 1.90 (1.56, 2.26) | 2 |
| Chad | 1.03 (1.01, 1.08) | 1 |
| Comoros | 2.13 (1.81, 2.39) | 1 |
| Congo (Republic of) | 2.91 (2.31, 3.65) | 3 |
| Cote d'Ivoire | 1.69 (1.51, 1.86) | 4 |
| Democratic Republic of the Congo | 1.41 (1.15, 1.64) | 3 |
| Djibouti | 1.05 (1.01, 1.13) | 2 |
| Equatorial Guinea | 3.59 (3.27, 3.79) | 1 |
| Eritrea | 3.01 (1.94, 3.79) | 1 |
| Ethiopia | 1.33 (1.19, 1.48) | 4 |
| Gabon | 3.34 (2.63, 3.79) | 1 |
| Gambia | 1.62 (1.39, 1.85) | 4 |
| Ghana | 1.78 (1.67, 1.90) | 7 |
| Guinea | 1.51 (1.28, 1.75) | 4 |
| Guinea-Bissau | 1.38 (1.01, 2.16) | 2 |
| Kenya | 2.26 (1.98, 2.58) | 5 |
| Liberia | 1.03 (1.01, 1.07) | 4 |
| Madagascar | 1.65 (1.48, 1.81) | 4 |
| Mali | 2.81 (2.46, 3.14) | 6 |
| Mauritania | 1.07 (1.01, 1.16) | 2 |
| Mozambique | 1.34 (1.21, 1.50) | 3 |
| Malawi | 1.33 (1.21, 1.45) | 9 |
| Niger | 3.50 (3.25, 3.78) | 2 |
| Nigeria | 2.22 (2.00, 2.47) | 9 |
| Rwanda | 1.59 (1.48, 1.70) | 6 |
| Senegal | 1.35 (1.22, 1.48) | 10 |
| Sierra Leone | 1.47 (1.31, 1.63) | 5 |
| Somalia | 2.35 (1.02, 3.66) | 1 |
| South Sudan | 1.02 (1.01, 1.04) | 3 |
| Sudan | 2.91 (2.11, 3.77) | 2 |
| Tanzania | 2.15 (1.88, 2.43) | 6 |
| Togo | 2.42 (2.21, 2.61) | 4 |
| Uganda | 1.66 (1.55, 1.78) | 6 |
| Zambia | 1.32 (1.21, 1.44) | 9 |
| Zimbabwe | 2.79 (2.26, 3.38) | 5 |

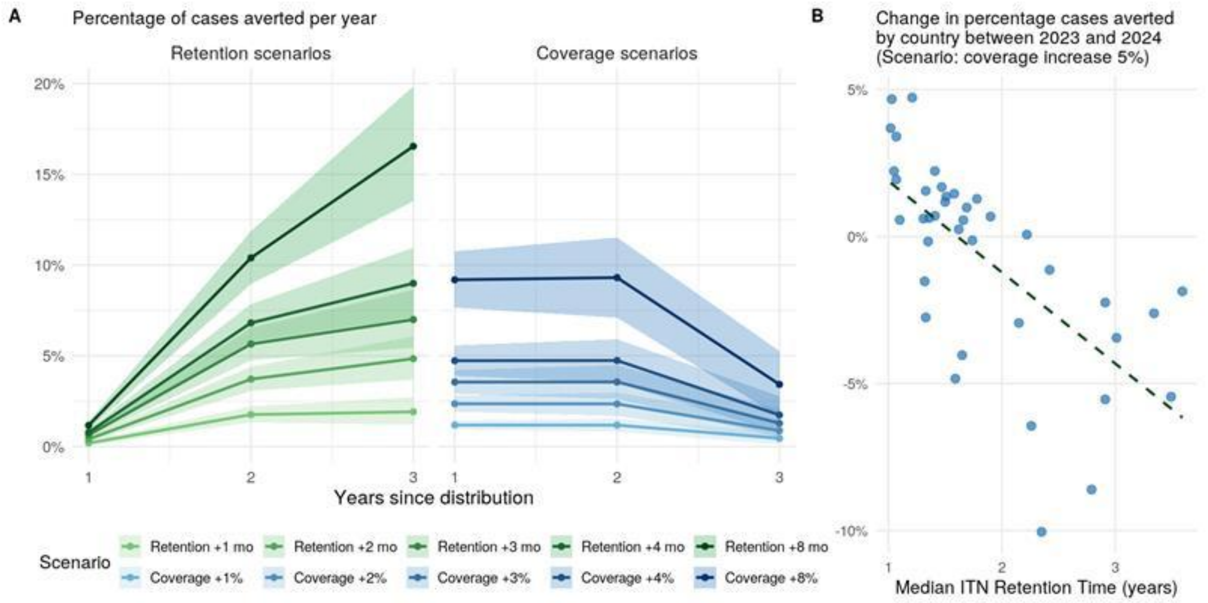


*Supplementary figure 1. A: Percentage of cases averted each year across settings in the increased retention (left) and increased coverage (right) scenarios. B: Country-level difference between percentage of cases averted in 2023 and 2024 plotted against baseline ITN retention times for the 5% increase in coverage scenario.*
